# Characterization and mitigation of aerosols and splatters from ultrasonic scalers

**DOI:** 10.1101/2021.02.26.21252487

**Authors:** Qisheng Ou, Rafael Grazzini Placucci, Judy Danielson, Gary Anderson, Paul Olin, Paul Jardine, John Madden, Qinghui Yuan, Timothy H. Grafe, Siyao Shao, Jiarong Hong, David Y.H. Pui

## Abstract

**Background:** Dental procedures often produce aerosols and splatter which have the potential to transmit pathogens such as SARS-CoV-2. The existing literature is limited.

**Methods:** Aerosols and splatter were generated from an ultrasonic scaling procedure on a dental mannequin and characterized by two optical imaging methods – digital inline holography (DIH) and laser sheet imaging (LSI). Capture efficiencies of various aerosol mitigation devices were evaluated and compared.

**Results:** The ultrasonic scaling procedure generates a wide size range of aerosols up to a few hundred micrometers and occasional large splatter which emit at low velocity (mostly below 3 m/s). Use of a saliva ejector (SE) and high-volume evacuator (HVE) resulted in 63% and 88% of overall reduction respectively while an extraoral local extractor (ELE) resulted in a reduction of 96% at the nominal design flow setting.

**Conclusions:** The study results showed that the use of ELE or HVE significantly reduced aerosol and splatter emission. The use of HVE generally requires an additional person to assist a hygienist, while an ELE can be operated “hands-free” when a dental hygienist is performing ultrasonic scaling and other operations.

**Practical Implications:** An extraoral local extractor aids in the reduction of aerosols and splatters during ultrasonic scaling procedures, potentially reducing transmission of oral or respiratory pathogens, like SARS-CoV-2. Position and airflow of the device are important to effective aerosol mitigation.

## Introduction

There is a general consensus that aerosols are one of the major paths of transmission for SARS-CoV-2 in the current pandemic^1^. This has led to concerns for health care workers involved in procedures which generate aerosols^2–5^. Dental providers are thought to be at particular risk due to the generation of aerosols and splatters^6–10^ during dental procedures such as ultrasonic cleaning and high-speed, water-cooled tooth preparation. The dental profession has responded with increased use of personal protection equipment (PPE) and recommendations to avoid aerosol generating procedures such as ultrasonic scaling. Current industry guidelines for addressing airborne risks focus on increased ventilation (or air changes per hour (ACH)), portable room filtration systems (not at the source), the avoidance of discretionary aerosol-generating procedures (AGPs), and adding time between patients and procedures to allow for increased ventilation^6,7,11^. These measures have understandably placed increased burden on the oral health care system and are based on limited data regarding the generation and mitigation of dental aerosols. Recommendations about mitigation of airborne contaminants in other occupational settings suggest that capture of contaminants near the source is far superior to general ventilation and/or use of PPE^12^.

There is a serious deficiency in the literature regarding the risks posed by aerosols and splatter from aerosol-generating procedures (AGPs) in dental settings and the efficacy of various aerosol mitigation techniques. A number of studies have collected aerosols and splatters directly onto a collecting surface for subsequent analysis, which include fluorescent^13–17^ or non-fluorescent^18–20^ based chromatic indicators and microbiological methods using culture media^21–24^. These studies are limited by their inefficient collection of small size aerosols (<∼50 µm) which do not provide a comprehensive characterization over the entire size spectrum. This is especially important as smaller aerosols may carry SARS-CoV-2 virus (0.06 −0.14 µm^25^). Some investigations have used aerosol sampling techniques^21,26–31^, but suffer from limited sampling of large aerosols and splatters or incorrect use of the sampling devices.

Dental procedures generate both aerosols and splatter. Without a commonly recognized size threshold, splatters are generally considered to be large droplets and/or debris that is heavy enough to settle rapidly without spreading a long distance. Aerosols, on the other hand, are small droplets that can become smaller through evaporation and result in smaller residual aerosols (10 microns in size or smaller) that may stay suspended in air indefinitely since they are sufficiently small to overcome gravitational settling, especially in a thermally stratified indoor environment^32–34^. Although most of these residual aerosols are the result of the supplied cooling water, they can include patient saliva, dental plaque, calculus, and blood, which can be infectious to dental practitioners or other dental patients if a patient carries SARS-CoV-2 or other infectious agents. Since a droplet’s ability to spread and subsequently be inhaled strongly depends on its original size and evaporation process, a comprehensive size characterization of the original droplets and the residual aerosols is essential in risk assessment of AGPs. Such a comprehensive assessment has not been reported yet to our best knowledge.

One example of a common dental hygiene procedure which is being avoided during the pandemic due to aerosol generation is ultrasonic scaling for dental prophylaxis^4^. This ultrasonic scaling technology increases the efficiency of dental hygienists and is much less physically demanding than manual scaling. Dental hygienists are at risk for repetitive stress injuries and the ban on ultrasonic scaling procedures makes this more likely. The reduced efficiency of manual scaling also increases the time the patient is seating in the dental chair and for some patients may increase the number of appointments needed. High-volume evacuation (HVE) is reported to reduce aerosol contamination considerably^23,35,36^, but it is generally not used by dental hygienists without an additional assistant to hold, position and manipulate the end of the HVE. Extraoral local extractors (ELE) have been discussed and marketed since the beginning of this pandemic, but the evidence supporting their effectiveness is rare^37,38^ and details about the proper design, configuration and use of ELEs has not been studied and reported.

We conducted this experiment to provide detailed physical characterizations of aerosols and splatters from ultrasonic scaling processes using novel *in-situ* optical methods. The methods were further used to evaluate the effectiveness of a variety of dental aerosol mitigation devices.

## Methods

Digital Inline Holography (DIH) and Laser Sheet Imaging (LSI) were used to characterize the size and velocity of aerosols and splatters from an ultrasonic scaling procedure. DIH can be leveraged to image particles larger than 6 μm but is limited to a small sample volume. On the other hand, LSI is not capable of distinguishing particle sizes, but can be used to count particles in a wide field of view. The combination of DIH and LSI measurements was therefore employed for more holistic insight into the generation and mitigation of aerosols during the dental procedure. This also provided an effective methodology for the evaluation of various aerosol mitigation strategies and devices.

### Digital Inline Holography (DIH)

Near-field *in situ* holographic measurements were conducted using a DIH setup consisting of a laser, a digital camera, as well as beam expanding, collimating and condensing optics. A dental mannequin with thermoplastic teeth was placed on a horizontal surface facing upwards to mimic a patient during the dental procedure. The center of the DIH sample volume (15.6×13.0×250 mm^3^) was located approximately 8 mm above the mannequin’s mouth opening (Figure 1).

**Figure 1.**
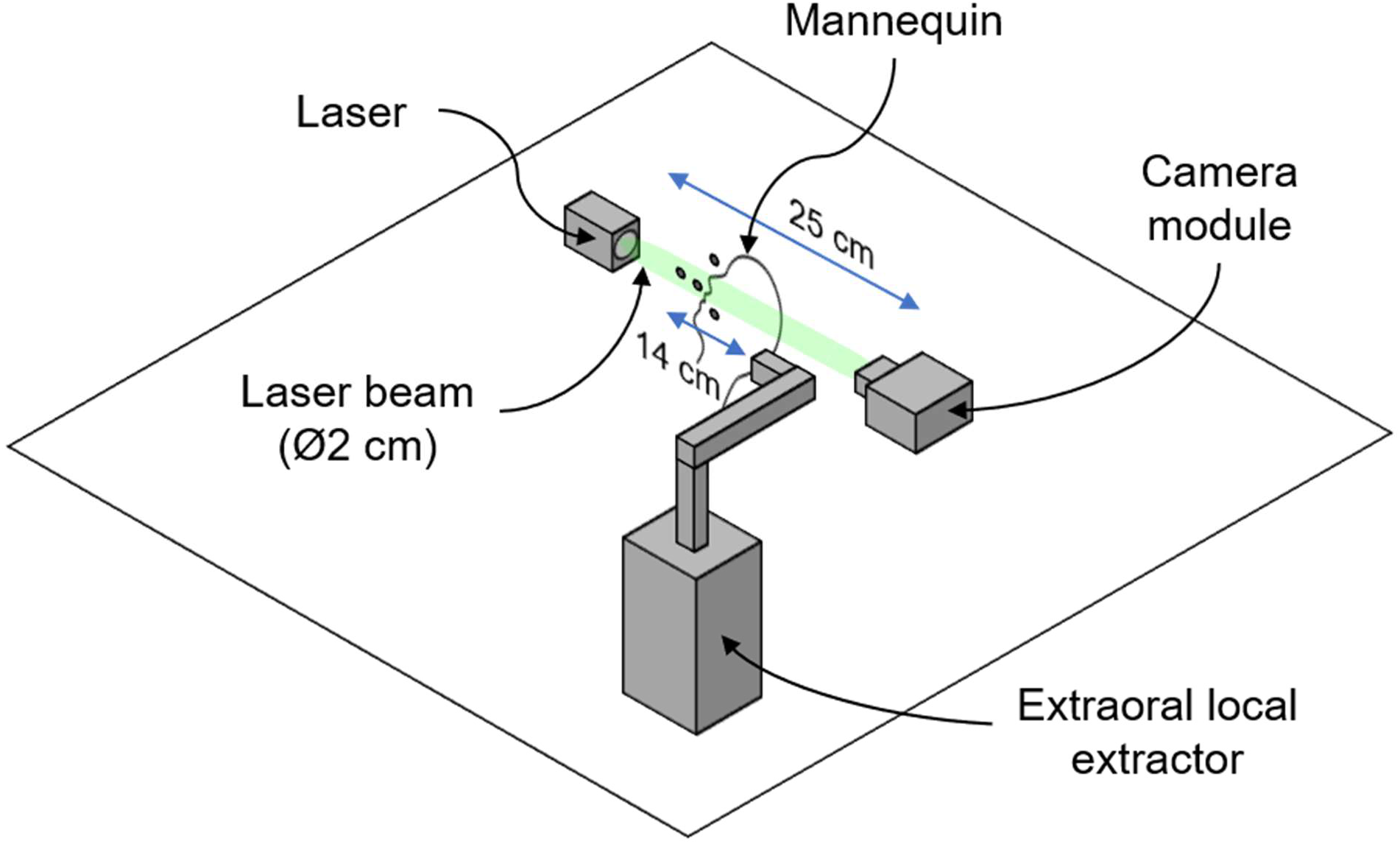
Schematic of DIH setup used for near-field aerosol size and velocity measurements. The DIH system is mounted onto a cage system secured to a table (not shown) and incorporates beam expanding, collimating and condensing optics (also not shown). A detailed description of the system can be found in Supplementary Information.

The hybrid hologram processing method proposed by Shao et al.^39^, consisting of image enhancement, digital reconstruction, particle segmentation and post-processing was used to obtain the size distribution of particles passing through the DIH sample volume during simulated dental procedures.

### Simulated Ultrasonic Scaling

A dental hygienist simulated supragingival scaling of the facial surfaces of the upper incisors and the lingual surfaces of the lower incisors using an ultrasonic scaler (Dentsply Sirona Cavitron ® Plus with 30K FSI-1000 inserts) operating at the medium power setting while the holographic sensor (35 fps, 13 μs exposure) acquired 10-second videos for a total of three trials. Clinically ultrasonic scaling in these dental regions generates the most splatters and aerosols.

### Tested Mitigation Devices and Strategies

Various mitigation devices were subsequently tested including a saliva ejector (SE, operated by the hygienist), a high-volume evacuation (HVE, operated by a second person) and an extraoral local extractor (ELE, BOFA DentalPro Aerosol UVC) at two different flow rates, 100 m^3^/h and 220 m^3^/h. Different aerosol mitigation devices, and combinations thereof, were employed during the simulated procedure. For the mitigation measurements the saliva ejector and HVE were used adjacent to the operating field as is typical of clinical practice. The ELE nozzle was tested at two distances from the mouth of the mannequin, 14 cm and 18 cm. The 14 cm distance was felt to be the most effective nozzle placement based on the recommendations of dental hygienists who collaborated with the manufacturer of the tested ELE through informal clinical testing. The 18 cm distance was investigated to further assess the impact of ELE nozzle position on aerosol mitigation.

### Laser Sheet Imaging (LSI)

Far-field *in situ* laser sheet imaging was conducted using the setup illustrated in Figure 2, which comprised of a high-resolution camera coupled with an image intensifier and an imaging lens, as well as a light sheet generating system. The dental mannequin was placed in a horizontal surface facing upwards in a fashion similar to the previous setup. The light sheet generating system was placed 80 cm to the side of the center of the camera’s field of view (FOV) and used to generate a light sheet that crossed the mannequin’s head just above its mouth opening and below its nose. The camera module, located 60 cm away from the mannequin, was oriented perpendicularly to the light sheet and focused to achieve a FOV of 40×40 cm^2^. Additional measures were taken to ensure the signal to noise ratio was maximized, such as spray-painting the mannequin head and implementing a light trap to diminish background reflection.

**Figure 2.**
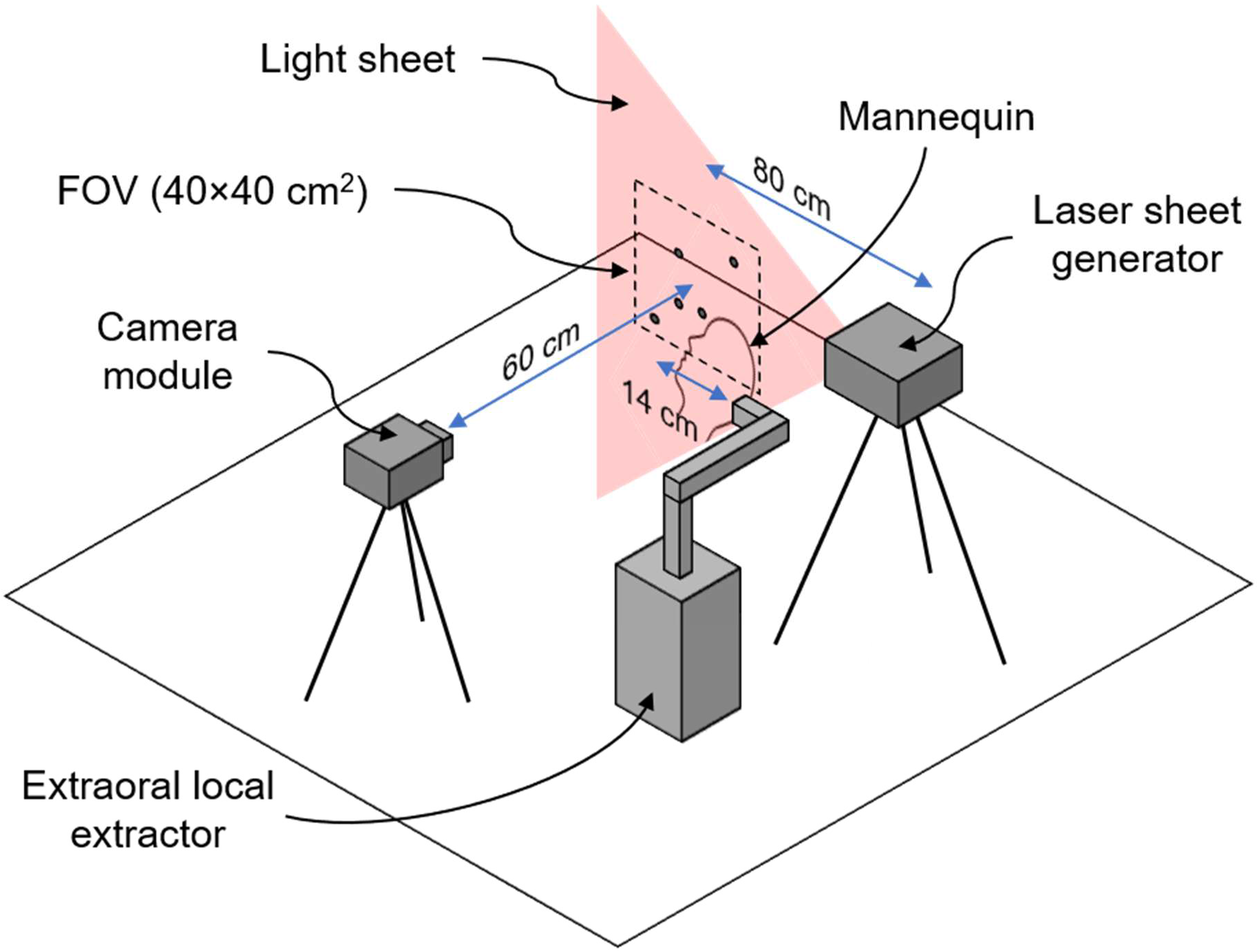
Schematic of LSI setup used for far-field plume and splatter measurements. The camera module incorporates a camera sensor, image intensifier and imaging lens, and the laser sheet generator is composed of a high-power laser and a set of optical lenses. A detailed description of the system can be found in Supplementary Information.

Recorded images were converted grayscale and enhanced using background subtraction. The plume of aerosol generated during the procedure was segmented by employing an entropy filter, and the total pixel intensity of the plume area was measured and compared to the mitigation-free test trial to obtain the device’s capture efficiency.

The ultrasonic scaling simulation and the mitigation techniques were tested with LSI in the same manner previously described with DIH. This provided a complementary broader field of view for the generated droplets and aerosols, as well as their mitigation.

## Results

### Digital Inline Holography (DIH)

#### Droplet size distribution

The observed droplet generation of the ultrasonic scaler was very dynamic consisting of a relatively continuous generation of a plume of small droplets (aerosols) with occasional large splatters shooting out at high speed. Like many other liquid-films broken up by mechanical forces, a wide range of droplet sizes was observed with DIH with more than 99% falling between 12 to 200 µm (Figure 3). Splatters larger than 200 µm were much less prevalent and were highly dependent on the positioning and movement of the ultrasonic scaler tip on teeth. The number mode (size with the highest number concentration) of the size distributions were 55 µm when working on the facial surfaces of teeth #8 & 9 and 34 µm for the lingual surfaces of teeth #24 & 25). Total droplet concentration was 114 particles per cubic centimeter (CC) for teeth #8 and 9 and 42 particles/CC for teeth #24 & 25. The lower concentration for teeth #24 and 25 was due to the more confined space trapping droplets inside the mouth of the mannequin. The concentration difference between the two sites was more pronounced for larger size droplets.

**Figure 3.**
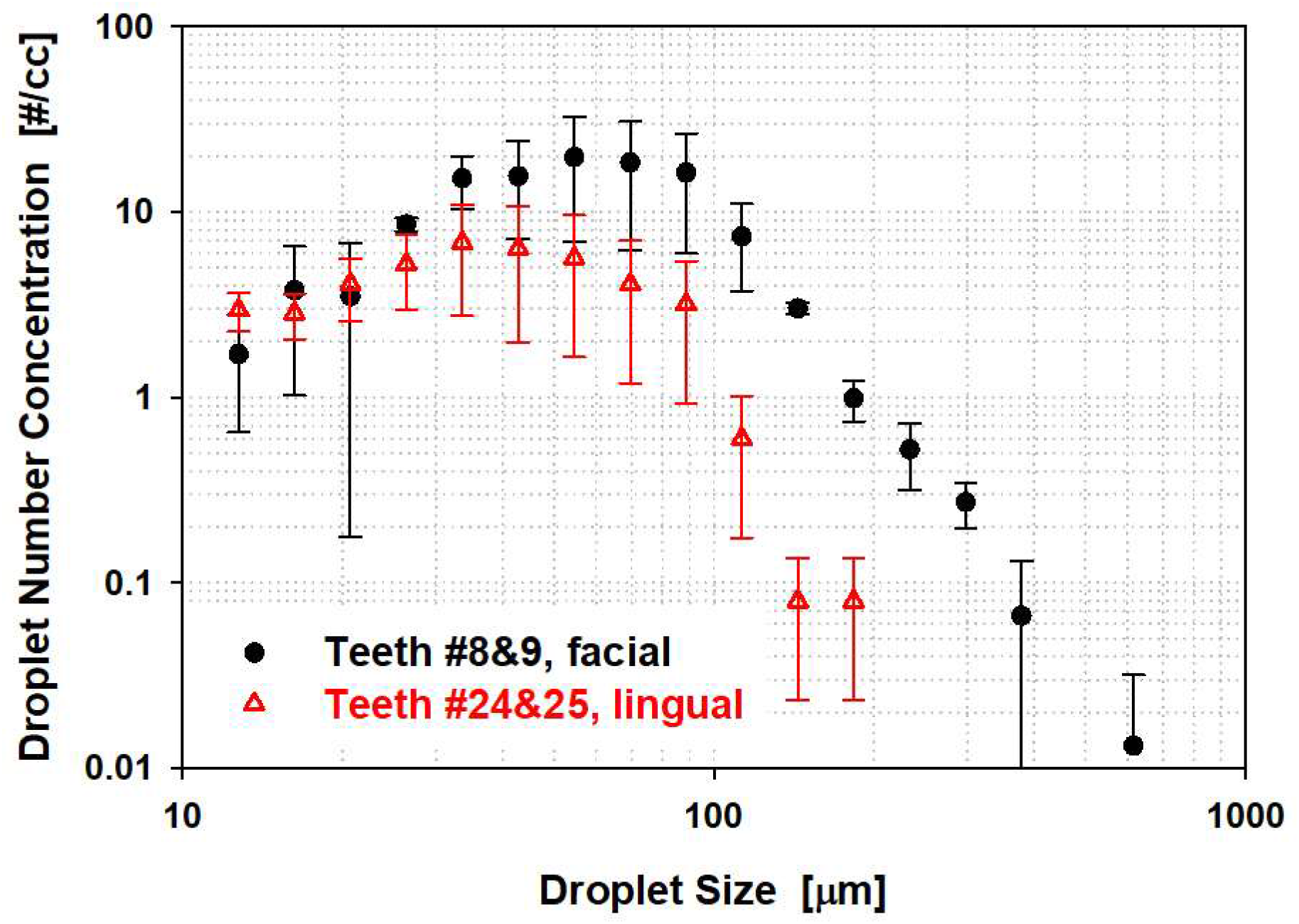
Size distribution of droplets as measured by DIH. (Error bars represent standard deviations of three measurements.)

#### Droplet velocity

More than 65% of the droplets had a velocity below 1 m/s and less than 2% of the droplets had a velocity greater than 3 m/s (See Figure 4). The low average droplet velocity observed here suggests a suction-based point-of-source mitigation strategy is possible and may be more effective than a whole-room-ventilation-based mitigation. A 3-D mapping of the velocity profile (not shown) suggested there was no dominant direction of droplet travel.

**Figure 4.**
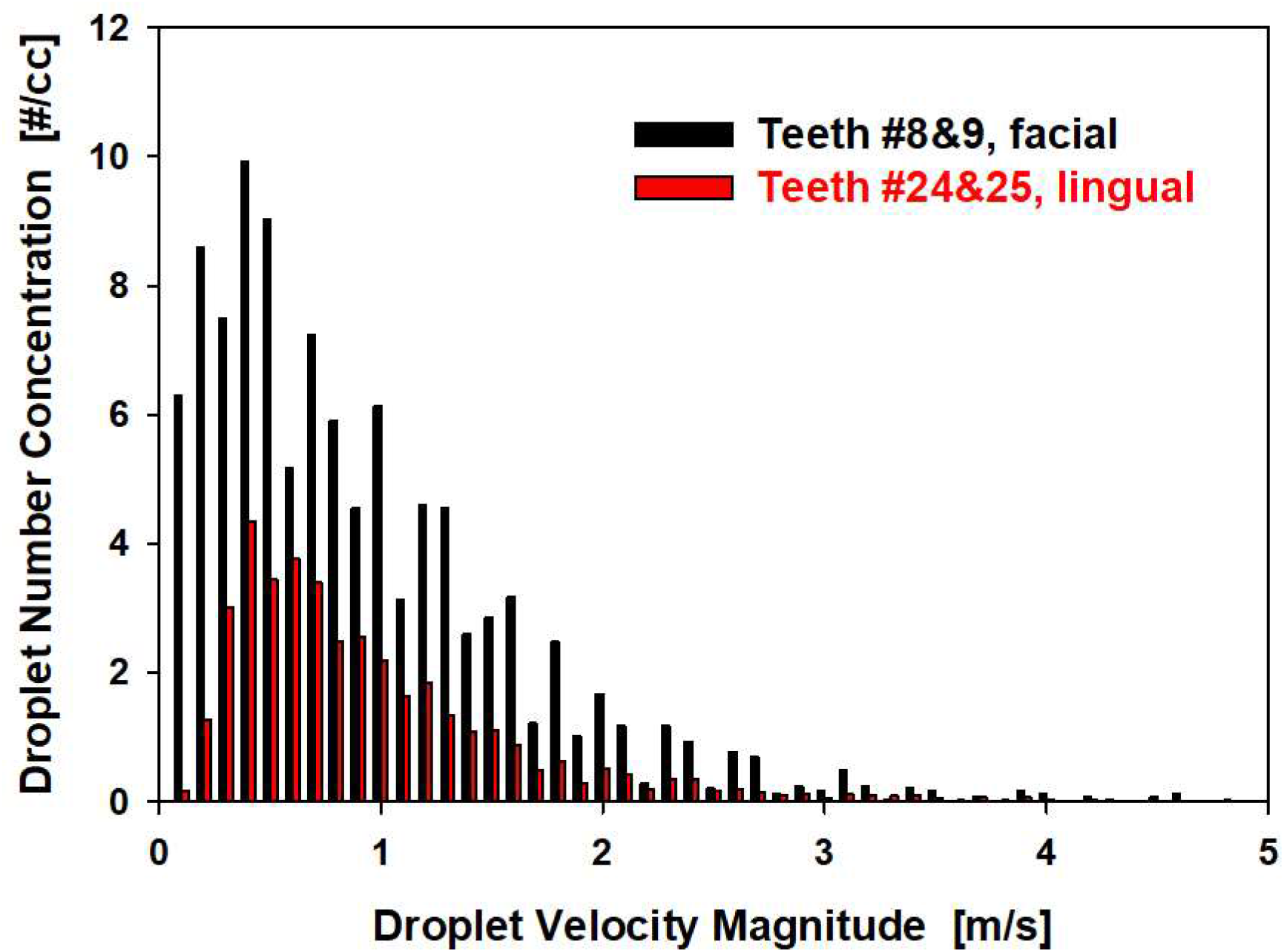
The velocity distribution of droplets as measured by DIH

#### Capture efficiency of mitigation devices

The capture efficiency of various mitigation devices is presented in Figure 5. The size dependent capture efficiency was determined by comparing the measured droplet size distribution with the mitigation device(s) applied with that measured when no mitigation was applied. There was a clear trend of decreased capture efficiency with increased droplet size. For the particular field of view of the DIH in this experiment, all mitigation devices but one (saliva ejector) demonstrated greater than 95% of capture for droplets up to 90 µm. The capture efficiency dropped to ∼ 80% and ∼ 50% at 120 and 150 µm, respectively. The previously described irregular generation of large splatters made a statistically meaningful quantification of capture efficiency impossible at droplet size larger than 180 µm. As previously discussed, droplets and aerosols smaller than 150 µm are the focus of any mitigation strategies given their high potential for longer-distance travel and transmission of SARS-CoV-2 and other airborne infectious agents.

**Figure 5.**
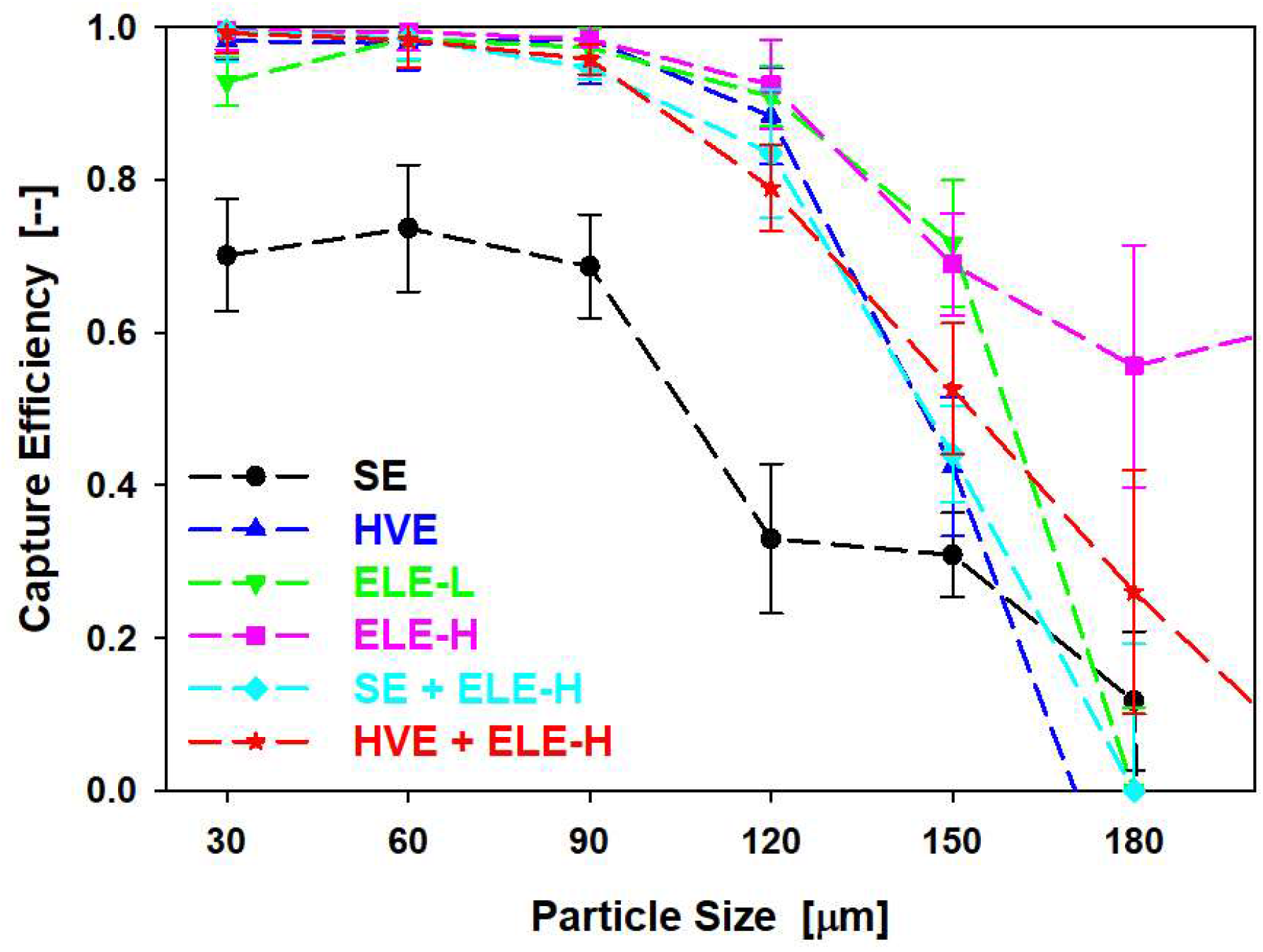
Size dependent capture efficiency of various mitigation device(s) by DIH when cleaning facial surface of teeth #8 & 9. (SE: saliva ejector; HVE: high-volume evacuation; ELE-L: extraoral local extractor at low flow setting; ELE-H: local extractor at high flow setting; ‘+’ denotes a combination of two devices. Error bars represent standard deviations of three measurements.)

### Laser Sheet Imaging (LSI)

LSI provided a larger field assessment of the tested mitigation strategies with more comprehensive visualization of their effect on aerosol and splatter spread. A sample video clip of LSI is provided in supplementary information, in which a local extractor was off first and turned on for ∼ 10 seconds and then turned off again to demonstrate its effectiveness. When no mitigation device was deployed, the plume (mainly clusters of small droplets at low speed) shows a swirling movement representing the vortexing flow generated by an ultrasonic scaler tip moving at high frequency. Larger splatters were generated occasionally (larger and brighter dots in the video) shooting out at higher speed. When the local extractor was turned on, the signal of both the plume and the distinct splatters were significantly suppressed, with a clear sign of the plume being drawn towards the opening of the extractor. Occasional splatters were still observed, consistent with the size-dependent capture observed by DIH that droplets with large momentum (larger size and/or higher speed) have relatively higher chance to escape from the capture of a mitigation device. Without distinguishing individual particles and their sizes, an overall capture efficiency was obtained from LSI data for each mitigation device (or their combinations) as shown in Figure 6. Among saliva ejector, HVE, and ELE at 14 cm working distance, saliva ejector had the lowest capture efficiency at 63%, followed up the local extractor at low flow setting (74%). HVE showed a higher capture efficiency at 88%. The ELE operating at high speed setting provided the best capture with an efficiency of 96%. A combination of local extractor at high flow setting with either saliva ejector (94%) or HVE (95%) did not provide additional improvement in capture efficiency, which implies the best mitigation strategy may be to use a local extractor (with proper flow setting, nozzle design, and placement relative to the patient and their oral cavity).

**Figure 6.**
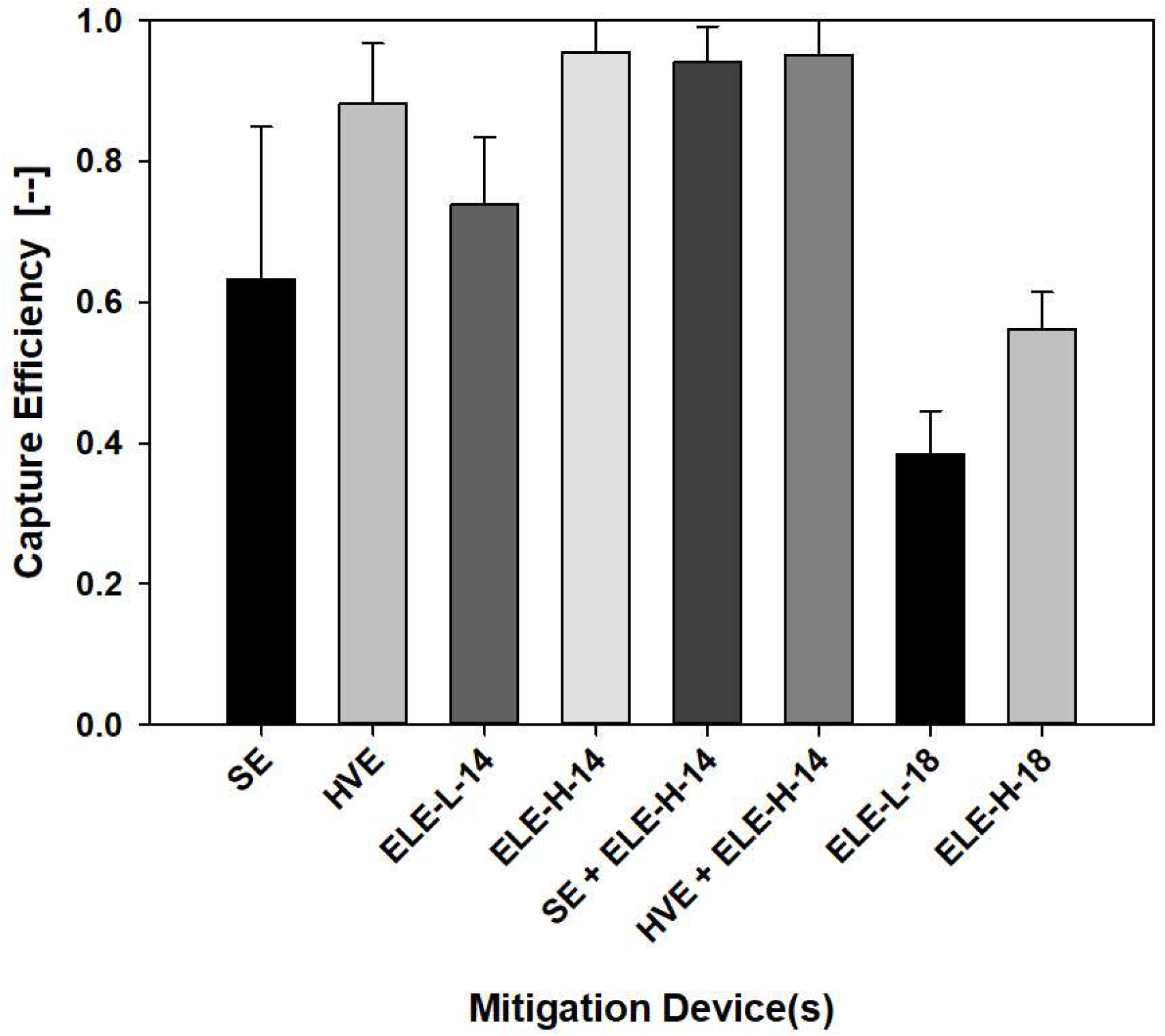
Capture efficiency of various mitigation device(s) by LSI when cleaning facial surface of teeth #8 & 9. (SE: saliva ejector; HVE: high-volume evacuation; ELE-L-#: local extractor at low flow setting at a distance of # cm; ELE-H-#: local extractor at high flow setting at a distance of # cm; ‘+’ denotes a combination of two devices. Error bars represent standard deviations of three measurements.)

The reported efficiencies for LSI were consistently lower than those by DIH at size from 30 to 120 µm, which can be explained by two reasons: (1) the existence of large splatters (with lower capture efficiency) in LSI video reduces the overall efficiency since a single large splatter contributes more intensity signal than a small droplet; and (2) the differences in size and position of the field of view between DIH and LSI result in characterizations of different portions of aerosol and splatter populations from the same source.

When the local extractor was moved 4 cm further away (from 14 to 18 cm) from the generation site, its capture efficiency dropped substantially from 74% to 38% at low flow setting, and from 96% to 56% at high flow setting.

## Discussion

Dental aerosols and splatters are an important potential transmission mode for many pathogens, including SARS-CoV-2, and it is vital to understand their generation profile and transportation behaviors in air to support the risk assessment and aerosol mitigation in the current COVID-19 pandemic. The combination of the DIH and broader field of LSI provided important complementary characterization of aerosols generated by an ultrasonic scaling with and without mitigation.

### Ultrasonic Scaler Aerosol Characterization

Among various engineering parameters, the size and velocity of original droplets are the most important ones that influence how they spread, evaporate, and their ability of being inhaled to cause potential aerosol transmission. To our best knowledge, this study characterizes these two important droplet properties for the first-time using novel *in-situ* optical methods. We demonstrate the wide spread of droplet sizes and the presence of both small and large droplets in the population. Small aerosols (<∼50 µm) are not sampled effectively by any impingement-based collection surface especially when their velocity is low due to the lack of momentum in air. Our findings suggest these small aerosols likely have been overlooked in previous studies employing surface collection methods for droplet sampling.

The droplet size and velocity reported here are properties measured close to their generation site outside of the oral cavity. When they travel further in the air, the droplets evaporate to significantly smaller size and the velocity relaxes close to the movement of the surrounding air. In view of the respiratory disease transmission, particles smaller than 10 µm (the size after droplets have reduced in size due to evaporation) pose more risk than larger ones since they are more likely to remain suspended in air^40^, to bypass a face shield, to penetrate through face masks / respirators^41–43^, and be inhaled and deposited in the respiratory tract. The residual particle size depends on the size and composition of the original droplets. For dental generated droplets, especially ultrasonic scaling, this is dominated by dissolved solid impurities in the cooling water which is typically tap water filtered with a bacterial filter. Assuming a total dissolved solid (TDS) level of 350 ppm for a typical tap water^44,45^, a droplet with a 142-µm original size dries into a 10 µm solid residual, suggesting droplets smaller than ∼150 µm should receive more attention in dental aerosol mitigation. Larger droplets, however, can deposit onto ground, dental chairs and other equipment, bodies of patients or dental operators, surfaces of face shield, face mask, or respirators, causing potential surface contamination. These findings therefore support the continued use of enhanced PPE in addition to regular surface cleaning and disinfection. The optical methods used in this study focus on initial droplets greater than 12 µm since any droplets smaller can be effectively mitigated as suggested by the size-dependent capture efficiency shown in Figure 5.

### Aerosol Mitigation

The ultrasonic scaler did produce omnidirectional droplets as expected, due to the high-frequency oscillation of the ultrasonic tip. However, aerosol mitigation was demonstrated and the ELE was very effective as compared to the more traditional clinical mitigation strategies without the need for additional personnel. It was clear that proper positioning of the nozzle and airflow of the ELE are of great importance but could be accomplished without impeding the work position of the dental hygienist.

The low average droplet velocity observed in this study suggests suction-based point-of-source mitigation strategies such as those tested may be more effective than whole-room-ventilation-based mitigation as has been recommended in other settings^12,46^. The ELE works in a similar principle as a saliva ejector or an HVE, but at a higher flowrate and a longer working distance. Unlike an HVE, it can be designed to remain in a fixed position, so an extra assistant is not needed to operate it.

It should be noted that room ventilation provided by heating, ventilation and air conditioning systems (HVAC) alone would require significantly higher flow for aerosol mitigation due to lack of focus on contamination sources. And it is possible that air ventilation provided by an HVAC system could contribute to aerosol spread rather than mitigation, as an aerosol could travel throughout an entire room before eventually being vented by the room HVAC system (where it may be recirculated if not properly filtered.) With ELE, however, the majority of contaminants are captured by the ELE at the source and subsequently filtered.

### Dental Aerosol and Infection Risk

Not every aerosol or splatter from ultrasonic scalers carries pathogens, so the emission profiles measured in this study represent a worst-case scenario, which need to be combined with a biological property evaluation for a more comprehensive risk assessment in the future. Since our knowledge on the infective dose of SARS-CoV-2 required to cause COVID-19 is still limited, it is difficult to draw definitive conclusions as to the risks posed by dental aerosols and splatters and if the reported efficacy of the mitigation devices is sufficient. It is therefore recommended that multiple mitigation strategies be employed to minimize the risk of dental aerosol transmission. The authors also recommend that dental practitioners continue to use enhanced PPE in addition to other aerosol mitigation techniques.

## Conclusions

Results of this study provide a scientific basis for risk assessment of aerosols and splatters from ultrasonic scaling. The engineering approaches of size and velocity profile characterization offer insight from a different angle – small aerosols which may be overlooked by a good number of previous studies can pose higher risk in terms of infection disease transmission through aerosol route, especially with the current COVID-19 pandemic. Implementing a hands-free extraoral local extractor can assist dental hygienists in implementing an effective, ergonomically sound method to reduce dental aerosols and splatters. Further research is needed on the biological property of these emissions for a comprehensive risk assessment. The methods in this work can be applied in the future to characterization of other AGPs in dental and medical settings to develop safe and efficient clinical practices in the face of highly contagious air-borne diseases. Future investigations will include assessment of water-cooled high-speed handpieces, as well as the interplay of point of source mitigation and clinical HVAC systems.

## Supporting information

Media file 1

Media file 2

## Data Availability

The authors confirm that the data supporting the findings of this study are available within the article and its supplementary materials.

## References

1. Karia R, Gupta I, Khandait H, Yadav A, Yadav A. COVID-19 and its Modes of Transmission. SN Compr Clin Med. 2020;2(10):1798–1801. doi:10.1007/s42399-020-00498-4

2. Harding H, Broom A, Broom J. Aerosol-generating procedures and infective risk to healthcare workers from SARS-CoV-2: the limits of the evidence. J Hosp Infect. 2020;105(4):717–725. doi:10.1016/j.jhin.2020.05.037

3. Guo W, Chan BH, Chng CK, Shi AH. Two Cases of Inadvertent Dental Aerosol Exposure to COVID-19 Patients. Ann Acad Med Singapore. 2020;49(7):514–516. doi:10.47102/annals-acadmedsg.2020186

4. Ge Z, Yang L, Xia J, Fu X, Zhang Y. Possible aerosol transmission of COVID-19 and special precautions in dentistry. J Zhejiang Univ Sci B. 2020;21(5):361–368. doi:10.1631/jzus.B2010010

5. Mick P, Murphy R. Aerosol-generating otolaryngology procedures and the need for enhanced PPE during the COVID-19 pandemic: A literature review. J Otolaryngol - Head Neck Surg. 2020;49(1):1–10. doi:10.1186/s40463-020-00424-7

6. World Health Organization. Considerations for the Provision of Essential Oral Health Services in the Context of COVID-19: Interim Guidance, 3 August 2020. World Health Organization; 2020.

7. Centers for Disease Control and Prevention. Interim Infection Prevention and Control Guidance for Dental Settings During the Coronavirus Disease 2019 (COVID-19) Pandemic. Published 2020. https://www.cdc.gov/coronavirus/2019-ncov/hcp/dental-settings.html#section-1

8. S KN, Eachempati P, Paisi M, Nasser M, Sivaramakrishnan G, Jh V. dental procedures for preventing infectious diseases (Review). Published online 2020. doi:10.1002/14651858.CD013686.pub2.www.cochranelibrary.com

9. Peng X, Xu X, Li Y, Cheng L, Zhou X, Ren B. Transmission routes of 2019-nCoV and controls in dental practice. Int J Oral Sci. 2020;12(1):9. doi:10.1038/s41368-020-0075-9

10. Amato A, Caggiano M, Amato M, Moccia G, Capunzo M, De Caro F. Infection control in dental practice during the covid-19 pandemic. Int J Environ Res Public Health. 2020;17(13):1–12. doi:10.3390/ijerph17134769

11. Office of Chief Dental Officer England. Standard operating procedure. Transition to recovery. 2020;(June):2–57. https://content/uploads/sites/52/2020/06/C0575-dental-transition-to-recovery-SOP-4June.pdf

12. The American Conference of Governmental Industrial Hygienists. Industrial Ventilation: A Manual of Recommended Practice for Design. 28th ed.; 2013.

13. Allison JR, Currie CC, Edwards DC, et al. Evaluating aerosol and splatter following dental procedures: Addressing new challenges for oral health care and rehabilitation. J Oral Rehabil. 2020;(August 2020):61–72. doi:10.1111/joor.13098

14. Dahlke WO, Cottam MR, Herring MC, Leavitt JM, Ditmyer MM, Walker RS. Evaluation of the spatter-reduction effectiveness of two dry-field isolation techniques. J Am Dent Assoc. 2012;143(11):1199–1204. doi:10.14219/jada.archive.2012.0064

15. Kaufmann M, Solderer A, Gubler A, Wegehaupt FJ, Attin T, Schmidlin PR. Quantitative measurements of aerosols from air-polishing and ultrasonic devices: (How) can we protect ourselves? PLoS One. 2020;15(12 December):1-10. doi:10.1371/journal.pone.0244020

16. Holliday R, Allison JR, Currie CC, et al. Evaluating contaminated dental aerosol and splatter in an open plan clinic environment: Implications for the COVID-19 pandemic. J Dent. 2021;105:103565. doi:10.1016/j.jdent.2020.103565

17. Harrel SK, Barnes JB, Rivera-Hidalgo F. Aerosol and Splatter Contamination from the Operative Site during Ultrasonic Scaling. J Am Dent Assoc. 1998;129(9):1241–1249. doi:10.14219/jada.archive.1998.0421

18. Shahdad S, Patel T, Hindocha A, et al. The efficacy of an extraoral scavenging device on reduction of splatter contamination during dental aerosol generating procedures: an exploratory study. Br Dent J. Published online 2020:1–10. doi:10.1038/s41415-020-2112-7

19. Harrel SK. Clinical use of an aerosol-reduction device with an ultrasonic scaler. Compend Contin Educ Dent. 1996;17(12):1185-1193; quiz 1194.

20. Chiramana S, O S, Kadiyala K, Prakash M, Prasad T, Chaitanya S. Evaluation of Minimum Required Safe Distance between Two Consecutive Dental Chairs for Optimal Asepsis. J Orofac Res. 2013;3:12–15. doi:10.5005/jp-journals-10026-1056

21. Zemouri C, Volgenant CMC, Buijs MJ, et al. Dental aerosols: microbial composition and spatial distribution. J Oral Microbiol. 2020;12(1). doi:10.1080/20002297.2020.1762040

22. Holloman JL, Mauriello SM, Pimenta L, Arnold RR. Comparison of suction device with saliva ejector for aerosol and spatter reduction during ultrasonic scaling. J Am Dent Assoc. 2015;146(1):27–33. doi:10.1016/j.adaj.2014.10.001

23. Bentley CD, Burkhart NW, Crawford JJ. Evaluating spatter and aerosol contamination during dental procedures. J Am Dent Assoc. 1994;125(5):579–584. doi:10.14219/jada.archive.1994.0093

24. Timmerman MF, Menso L, Steinfort J, Van Winkelhoff AJ, Van Der Weijden GA. Atmospheric contamination during ultrasonic scaling. J Clin Periodontol. 2004;31(6):458–462. doi:10.1111/j.1600-051X.2004.00511.x

25. Zhu N, Zhang D, Wang W, et al. A Novel Coronavirus from Patients with Pneumonia in China, 2019. N Engl J Med. 2020;382(8):727–733. doi:10.1056/NEJMoa2001017

26. Dutil S, Mériaux A, De Latrémoille MC, Lazure L, Barbeau J, Duchaine C. Measurement of airborne bacteria and endotoxin generated during dental cleaning. J Occup Environ Hyg. 2008;6(2):121–130. doi:10.1080/15459620802633957

27. Miller RL. Characteristics of Blood-Containing Aerosols Generated by Common Powered Dental Instruments. Am Ind Hyg Assoc J. 1995;56(7):670–676. doi:10.1080/15428119591016683

28. Bennett AM, Fulford MR, Walker JT, Bradshaw DJ, Martin M V, Marsh PD. Microbial aerosols in general dental practice. Br Dent J. 2000;189(12):664–667. doi:10.1038/sj.bdj.4800859

29. Jacks ME. A laboratory comparison of evacuation devices on aerosol reduction. J Dent Hyg. 2002;76(3):202–206.

30. Matys J, Grzech-Leśniak K. Dental aerosol as a hazard risk for dental workers. Materials (Basel). 2020;13(22):1–13. doi:10.3390/ma13225109

31. Nulty A, Lefkaditis C, Zachrisson P, Van Tonder Q, Yar R. A clinical study measuring dental aerosols with and without a high-volume extraction device. Br Dent J. Published online 2020:1-8. doi:10.1038/s41415-020-2274-3

32. Liu F, Qian H, Zheng X, Song J, Cao G, Liu Z. Evaporation and dispersion of exhaled droplets in stratified environment. IOP Conf Ser Mater Sci Eng. 2019;609(4). doi:10.1088/1757-899X/609/4/042059

33. Liu F, Zhang C, Qian H, Zheng X, Nielsen P V. Direct or indirect exposure of exhaled contaminants in stratified environments using an integral model of an expiratory jet. Indoor Air. 2019;29(4):591–603. doi:10.1111/ina.12563

34. Zhou Q, Qian H, Ren H, Li Y, Nielsen P V. The lock-up phenomenon of exhaled flow in a stable thermally-stratified indoor environment. Build Environ. 2017;116:246–256. doi:10.1016/j.buildenv.2017.02.010

35. Sawhney A, Venugopal S, Girish Babu RJ, et al. Aerosols how dangerous they are in clinical practice. J Clin Diagnostic Res. 2015;9(4):52–57. doi:10.7860/JCDR/2015/12038.5835

36. Harrel SK, Barnes JB, Rivera-Hidalgo F. Reduction of Aerosols Produced by Ultrasonic Sealers. J Periodontol. 1996;67(1):28–32. doi:10.1902/jop.1996.67.1.28

37. Teanpaisan R, Taeporamaysamai M, Rattanachone P, Poldoung N, Srisintorn S. The usefulness of the modified extra-oral vacuum aspirator (EOVA) from household vacuum cleaner in reducing bacteria in dental aerosols. Int Dent J. 2001;51(6):413–416. doi:10.1002/j.1875-595X.2001.tb00853.x

38. Junevicius J, Surna A, Surna R. Effectiveness evaluation of different suction systems. Stomatologija. 2005;7(2):52–57.

39. Shao S, Li C, Hong J. A hybrid image processing method for measuring 3D bubble distribution using digital inline holography. Chem Eng Sci. 2019;207:929–941. doi:10.1016/j.ces.2019.07.009

40. Hinds WC. Aerosol technology: properties, behaviour, and measurement of airborne particles. Published online 1982. Accessed January 24, 2021. https://www.wiley.com/en-us/Aerosol+Technology%3A+Properties%2C+Behavior%2C+and+Measurement+of+Airborne+Particles%2C+2nd+Edition-p-9780471194101

41. Pei C, Ou Q, Kim SC, Chen S-C, Pui DYH. Alternative Face Masks Made of Common Materials for General Public: Fractional Filtration Efficiency and Breathability Perspective. Aerosol Air Qual Res. 2020;20(Viii):2581-2591. doi:10.4209/aaqr.2020.07.0423

42. Ou Q, Pei C, Chan Kim S, Abell E, Pui DYH. Evaluation of decontamination methods for commercial and alternative respirator and mask materials – view from filtration aspect. J Aerosol Sci. 2020;150:105609. doi:10.1016/j.jaerosci.2020.105609

43. Konda A, Prakash A, Moss GA, Schmoldt M, Grant GD, Guha S. Aerosol Filtration Efficiency of Common Fabrics Used in Respiratory Cloth Masks. ACS Nano. 2020;14(5):6339–6347. doi:10.1021/acsnano.0c03252

44. Park M, Snyder SA. Attenuation of contaminants of emerging concerns by nanofiltration membrane: rejection mechanism and application in water reuse. In: Contaminants of Emerging Concern in Water and Wastewater. Elsevier; 2020:177–206. doi:10.1016/B978-0-12-813561-7.00006-7

45. Quench - a Culligan company. What’s in Your Water: Total Dissolved Solids (TDS) in Drinking Water. Published 2019. https://quenchwater.com/blog/tds-in-drinking-water/

46. Li A, Kosonen R, Hagström K. Industrial ventilation design method. In: Industrial Ventilation Design Guidebook. ; 2020:19–37. doi:10.1016/b978-0-12-816780-9.00003-4

