## Supplementary Information for "Characterization and mitigation of aerosols and splatters from ultrasonic scalers"

### A1. Supplemental Material – Digital Inline Holography (DIH)

DIH utilizes a coherent light source (e.g., laser) to illuminate suspended particles (e.g., droplets, bubbles, dust, cells, etc.) in a three-dimensional sample volume, and a digital sensor (e.g., camera) that records the patterns (referred to as holograms hereafter) generated by the interference between light scattered from each object and the non-scattered portion of the illumination beam[1]. These holograms are subsequently reconstructed at several planes using different diffraction kernels to form a 3D optical field. The location, size and morphology of objects in the sample volume can then be extracted.

The DIH setup used to characterize the size and velocity distribution generated during the dental procedures is illustrated in Figure 1. A continuous-wave laser with 532 nm wavelength and 4.5 mW power is employed as the source of coherent illumination. A 40X microscopic objective is used to expand the beam to a diameter of approximately 2 cm, and a f=60 mm bi-convex lens is leveraged for beam collimation. The beam is condensed using a f=35.7 mm aspheric lens, and a 1:1.4/25mm imaging lens is utilized to reduce spherical distortion and achieve the desired magnification. Holograms are acquired using a digital camera with a 2448×2048 pixel$^2$ CMOS sensor operating at 35 Hz and an exposure time of 13 μs. All components secured onto a cage system which is mounted onto an optical rail (not shown). A 3D-printed case (not shown) is used to enclose all the optical components to minimize damage and contamination from the external environment. Altogether, this system features an image resolution of 6.34 μm/pixel (after magnification) and a sample volume of approximately 15.6×13.0×250 mm$^3$.

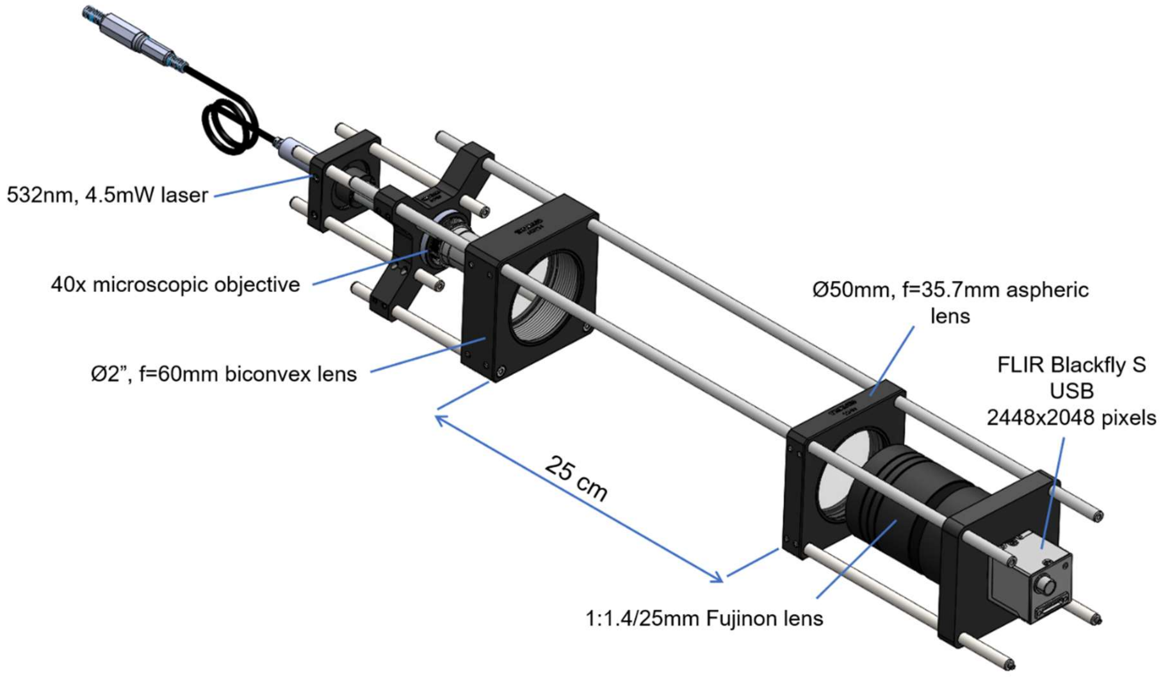


Figure 1: Schematic of DIH setup

**A2. Supplemental Material – DIH Sample Data**

Media file 1 illustrates a sequence of enhanced holograms (left) and the corresponding 3D rendering of the particles within the DIH sample volume (right). The particle field showcased in this video corresponds to a one second recording of a mitigation-free ultrasonic scaling procedure using a high-speed camera (1057 Hz, 5 μs exposure time). Particle sizes illustrated in the 3D render are augmented for better visualization and the field of view is equivalent to that discussed in §A1.

**A3. Supplemental Material – LSI Setup**

LSI employs a thin sheet of light to illuminate particles passing through it, which are observed by a sensor (e.g., camera). The position of particles crossing the light sheet are then extracted using digital image recognition algorithms and used to quantify other characteristics, namely particle trajectory, speed and concentration within the sample volume.

The laser sheet setup used to characterize the far-field behavior of aerosol and splatters generated during the dental procedures (shown in Figure 2) employs a low-light sensitivity camera

(Nikon D610), an image intensifier (AstroScope Gen 3), a macro lens (Nikkor 50 mm) and a laser sheet generating system. The laser sheet generating system consists of a high power continuous laser (638 nm, 2W), and a set of biconcave cylindrical lenses (f=10 mm and f=50 mm), and is capable of generating a 15 mm thick light sheet with a 24 degree half-angle. The camera is adjusted to capture full HD video (1920×1080 pixels$^2$) at 30 frames per second with an exposure time of 20 ms and ISO setting of 1000. The image intensifier is tuned to its maximum gain setting.

**A4. Supplemental Material – LSI Sample Data and Data Analysis**

Media file 2 is a video captured using the system described §A3 that demonstrates the ultrasonic scaling of the facial surface of teeth #8. The ELE unit was located 14 cm away from the mannequin's mouth and activated for 10 seconds at power level 3 approximately 10 seconds after the start of the video.

Following is a description of the data processing steps required to obtain the aerosol capture efficiency during the different dental procedures using LSI. The video acquired using the apparatus described in §A3 was first converted into a stack of grayscale images and enhanced by subtracting each individual frame by the mean intensity of the stack. The bottom and top 1% of all pixel values were saturated to increase the contrast of each frame. A binary mask was created to remove unwanted background artifacts (e.g., operator) and applied to the high-contrast image. The masked image was subsequently binarized using an adaptive thresholding function based on the local mean intensity in the neighborhood of each pixel. Larger particles (referred to as splatters herein) were segmented from the binarized image, and their pixel intensity was recorded.

For the analysis of concentrated aerosol (referred to as plume herein), the local entropy of the aforementioned high-contrast image stack was calculated, converted into a binary image, and used to create a mask to segment the plume boundary with the remained of the image. Once the plume was segmented, the intensity of each pixel within the plume region was recorded.

The pixel intensities for plume and splatter alike were summated and compared to that corresponding to the mitigation-free cases in order to obtain the device's capture efficiency. This entire procedure was repeated for several combinations of mitigation devices.

**Reference**

1. Katz J, Sheng J. Applications of holography in fluid mechanics and particle dynamics. *Annu Rev Fluid Mech*. 2010;42:531-555. doi:10.1146/annurev-fluid-121108-145508
